# Tongue Coating in COVID-19 Patients: A Case-Control Study

**DOI:** 10.1101/2022.03.14.22272342

**Authors:** Zhi Chun Wang, Xi Hong Cai, Jeremy Chan, Yi Yi Chan, Xiaotong Chen, Ching Wan Cheng, Donghui Huang, Bei-ni Lao, Xu-sheng Liu, Aiping Lu, Huixian Wang, Helen Zhang, Xuebin Zhang, Shi Ping Zhang

## Abstract

It has been suggested that COVID-19 patients have distinct tongue features, which may help to monitor the development of their condition. To determine if there was any specific tongue coating feature in COVID-19, this study investigated the difference in tongue coating between COVID-19 subjects and subjects with other acute inflammatory diseases characterized by fever. Tongue images taken with smartphones from three age-matched groups, namely, COVID group (n=92), non-COVID febrile group (n=92), and normal control group (n=92), were analyzed by two blinded raters according to a tongue coating scoring scheme, which assessed the levels of thick fur, slimy or greasy fur, discolored fur and composite index of tongue coating. Compared with control, significant increases in all coating indexes were found in the COVID group (P<0.001), as well as in the non-COVID febrile group (P<0.001). However, no difference was observed between COVID and non-COVID febrile groups for all coating indexes measured. In COVID-19 subjects, their scores of coating indexes had weak but significant correlations with certain inflammatory biomarkers, including WBC and neutrophil - lymphocyte ratio. It is concluded that COVID-19 subjects have pathological tongue coating patterns that are associated with inflammatory responses, and these coating patterns can help to indicate the direction of disease development.

## Introduction

COVID-19 is caused by infection with the severe acute respiratory syndrome coronavirus 2 (SARS-CoV-2) virus strain [1]. As an acute febrile illness, COVID-19 has led to severe health consequence worldwide. According to the World Health Organization, as of Febuary 2022, over 426 million cases have been confirmed, with more than 5.9 million confirmed deaths attributed to COVID-19, making it one of the deadliest pandemics in history [2]. It is axiomatic that symptom detection and monitoring are important in the fight against the disease.

One of the commonly seen symptoms of COVID-19 is the loss of taste sensation and a change in tongue appearance. An autopsy study with matched nasopharyngeal and saliva samples showed distinct virus shedding dynamics, and saliva viral load was associated with COVID-19 symptoms, including loss of taste [3]. It has been proposed that SARS-CoV-2 virus attacks the mucosa of oral cavity by targeting ACE2 receptors, which are highly enriched in epithelial cells of the tongue [4]. Early report from China suggested that certain specific tongue patterns such as tongue discoloration and pathological tongue coating were found in COVID-19 subjects [5-8]. A Spanish study of 666 COVID-19 patients found that half of the victims displayed symptoms of the tongue, including transient lingual papillitis, glossitis with lateral indentations, glossitis with patchy depapillation, burning sensation and taste disturbances [9]. It has been proposed that a specific type of tongue symptom, called “COVID tongue”, should be included into the list of common symptoms of COVID-19 [10].

However, the existence of COVID tongue has been questioned, with the argument that the changes of tongue appearance might be an anatomical variation of normality, rather than pathological [11]. Moreover, age is an important factor in tongue appearance [5]. However, in previous studies of COVID tongue [7-9], age matching has not been performed, exposing the results to bias.

Tongue coating changes had been proposed as a convenient sign for early detection of COVID-19 [8]. In our previous study, we evaluated the reliability of tongue diagnosis in traditional Chinese medicine (TCM) using images from smartphones, and found that the majority of tongue coating features, corresponding to different types of tongue fur in TCM, could be reliably represented in smartphone tongue images [12]. In view of the highly contagious nature of COVID-19, the possibility of using self-recorded tongue images obtained by smartphone to aid symptom detection and monitoring has practical appeals. Therefore, we carried out the present study with age matching to evaluate the differences in tongue coating features between COVID, non-COVID febrile and normal control subjects, based on tongue images captured with smartphones. The hypotheses to be tested was that there were significant differences in tongue coating features between COVID and non-COVID febrile subjects, and between COVID and normal subjects. Since the studied conditions involved inflammation, we also examined the correlations between tongue coating changes and inflammatory biomarkers.

## Methods

### Data collection

This study had obtained ethical approval from the relevant IRBs of institutions of contributing authors. Informed consent was obtained from healthy participants and non-COVID febrile subjects, but was waived for COVID subjects due to the retrospective nature of the study and the exceptional circumstances. A total of 478 images from COVID subjects, including their partial anonymous medical records, were provided by rapid response medical teams working in Wuhan during Feb-March 2020. After initial screening of their images and corresponding medical records, 141 patients were retained for further analysis. All the images from COVID patients were taken by smartphones with unknown methods. The included subjects had records of age, gender, diagnosis of COVID-19, and some had laboratory tests and outcomes of hospital stay, i.e., returning to home care or being transferred to another hospital for more intensive care.

For non-COVID febrile patients and control subjects, their tongue images were collected with a standardized smartphone photographic protocol using smartphones of common brand names, including iPhone, HUAWEI, and Samsung, as described in our previous paper [12]. The non-COVID febrile group was made-up of subjects from the Guangdong Province in the Third Affiliated Hospital of Guangzhou University of Chinese Medicine and the Guangdong Chinese Medicine Hospital Zhuhai Branch. These subjects had acute fever for various reasons but were tested negative for COVID-19. The normal control group consisted of subjects without any acute illness. To control for local diet, 51 subjects in the normal control group were derived from employees of a Wuhan catering company (Wuhan Xiao Pan Jing Hotel Management Co. Ltd), who underwent regular COVID-19 testing during the data collection period because of the nature of their job. The remaining control subjects (n=41), whose data were retrieved randomly from a tongue image database of our previous studies [12], were from Guangdong and Hong Kong.

As shown in Figure 1, the 92 subjects in COVID group were matched by subjects from the two other groups based on age (See Figure 1). There is no significant difference in gender composition between the three groups in the marched dataset (see Table 1).

**Table 3.**
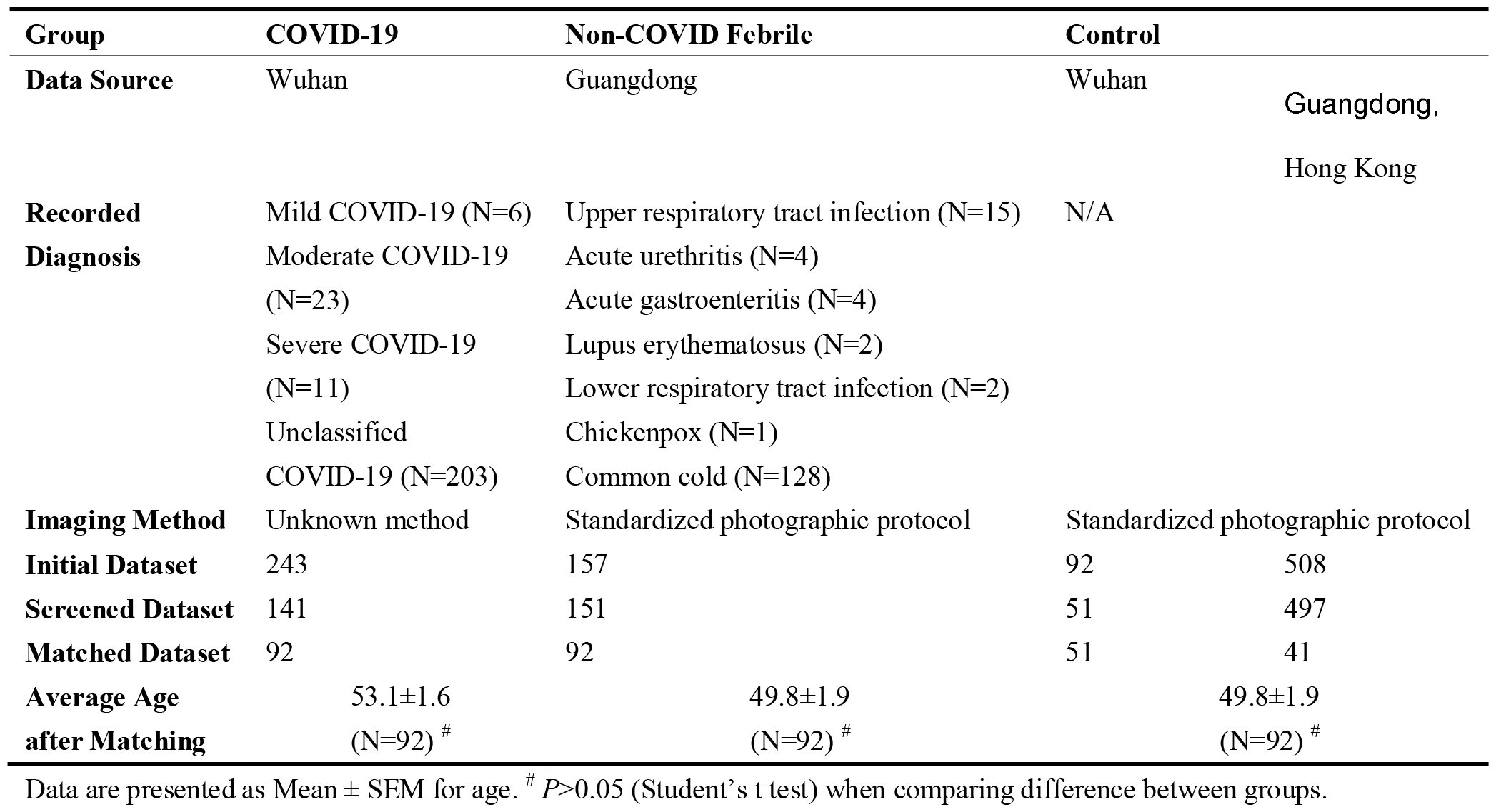
Data source and Baseline Information.

**Figure 2.** Flow-chart showing the process of subject matching for comparison of tongue coating

### Scoring for tongue coating indexes

Firstly, the color temperature of smartphone tongue images was adjusted using white balancing function of a commercial software (Adobe Photoshop CC 2019) to reduce the influence of different light sources. Then the method described by Winkel etc. [13] was used for scoring tongue coating indexes. Briefly, the tongue surface was divided into six areas (see Figure 2), and raters assigned three categories of scores to each area, quantifying the thickness of tongue coating (tongue coating index; TCI), the presence of slimy or greasy tongue fur (slimy biofilm index; SBI) and the presence of discoloration on tongue fur (tongue discoloration index; TDI). The scores from the six areas in each category were then added up respectively to become an index score for the tongue image. TCI as assessed by zooming in the photograph to observe individual papillae. SBI examined the presence of slimy biofilm, which was a white to whitish-yellow creamy confluent plaques (milk curd or cottage cheese-like). TDI indicated a change in color of tongue fur, which in normal subjects should be whitish or pink. The details of the scoring method are shown in Table 2. A composite tongue coating index (CTCI) was calculated by the sum of the three indexes. An example of the application of these indexes is shown in Figure 2.

**Table 2.** Rating Scheme for Tongue Fur Features at Each Subdivision.

| Tongue Fur Features | Score | Explanation |
| --- | --- | --- |
| Thick Fur | 0 | Normal thin coating or no coating |
|  | 1 | Thick coating, papillae of tongue just visible |
|  | 2 | Very thick coating, papillae of tongue not visible |
| Slimy Fur | 0 | Absence of slimy biofilm |
| | 1 | Slimy biofilm occupying $\leq 50\%$ of the area |
| | 2 | Slimy biofilm occupying $>50\%$ of the area |
| Fur discoloration | 0 | Pink color, or whitish color |
| | 1 | Other colors occupying $\leq 50\%$ of the area |
| | 2 | Other colors $>50\%$ of the area |

**Figure 2.** Smartphone tongue image from a COVID-19 subject (left) and diagram (right) showing an example of the subdivision and scoring schemes for calculation of tongue coating indexes. TCI: tongue coating index; SBI: slimy biofilm index; TDI: tongue discoloration index; and CTCI: composite tongue coating index.

Because there was no common standard in quantifying COVID tongue coating features, we conducted a pilot study on method agreement using six raters to evaluate a test set of COVID tongue images (n=100). The raters consisted of three Chinese medicine practitioners (CYC, YYC, BNL) and three Chinese medicine students (XTC, QYC, JC), who were blinded to the source and diagnosis of the images. We found that the interrater correlations on coating indexes between two raters ranged from 0.648 - 0.773 and identified two raters (YYC & JC) who had the highest correlation coefficient amongst all the combinations of pair. Since YYC was not available for further image assessment, we trained two student raters (QYC and JC) working together until their scoring of the test set reached a strong agreement (Pearson correlation = 0.762, *P*<0.001). The two raters then used the same method to score the final image sets of the three matched groups, which were mixed together with group status concealed. The average scores of their rating for each image were used for statistical analysis.

### Statistical analysis

Data were analyzed with IBM SPSS Statistics (version 26.0) and GraphPad Prism (version 8.0.2). Continuous variables were compared using the Student’s t test or the Mann-Whitney U test, depending on the distribution of the data. Correlation coefficient was calculated using Pearson correlation coefficient. Results with P < 0.05 are considered to be statistically significant.

## Results

### Comparison of tongue coating indexes between groups

Three age-matched groups of 92 subjects each were included in the comparative analysis (Table 1), with older age subjects in the COVID-19 group being excluded due to the lack of matching subjects. As seen in Table 3, the median scores of TCI, SBI, TDI and CITC of the COVID group and of the non-COVID febrile group are significantly higher than those of the control group (P<0.001). However, there was no significant difference in the median scores of TCI, SBI, TDI and CITC between the COVID group and the non-COVID febrile group.

**Table 3.** Comparison of Tongue Coating Indexes in Different Groups.

| Index | COVID<br>(N=92) | Non-COVID<br>Febrile (N=92) | Control<br>(N=92) | *P* between<br>COVID &<br>Non-COVID<br>Febrile | *P* between<br>COVID &<br>Control | *P* between<br>Non-COVID<br>Febrile &<br>Control |
| --- | --- | --- | --- | --- | --- | --- |
| TCI | 4.0 (2.0-6.0) | 4.3 (1.1-7.0) | 1.0 (0.0-2.0) | 0.484 | <0.001*** | <0.001*** |
| SBI | 2.0 (0.0-4.0) | 2.5 (0.6-5.0) | 0.0 (0.0-1.0) | 0.144 | <0.001*** | <0.001*** |
| TDI | 1.5 (0.0-4.0) | 2.0 (0.1-5.0) | 0.0 (0.0-1.0) | 0.197 | <0.001*** | <0.001*** |
| CTCI | 8.5 (3.0-13.0) | 9.5 (3.0-15.0) | 2.0 (0.0-5.8) | 0.253 | <0.001*** | <0.001*** |

Data are presented as Median (IQR). *P* values were calculated by Mann-Whitney U test. \* $P < 0.05$ , \*\* $P < 0.01$ , \*\*\* $P < 0.001$ .

### Comparison between tongue coating indexes with inflammatory biomarkers

We performed subgroup analysis in COVID-19 and non-COVID febrile patients of their tongue coating indexes and inflammatory biomarkers including C-reactive protein (CRP), white blood cell count (WBC), lymphocytes, hemoglobin, blood platelet count (BPC), neutrophils, neutrophils lymphocytes ratio (NLR) and platelet lymphocytes ratio (PLR). For COVID-19 patients, weak but significant correlations were observed between tongue coating indexes and WBC, neutrophils and NLR. For non-COVID febrile patients, only weak correlations were found between CRP and SBI, TDI and CTCI, with no correlation found between other biomarkers and tongue coating idexes (see Table 4).

**Table 4.** Correlation Coefficients between Tongue Coating Indexes and Biomarkers.

| Indexes | COVID-19 patients |  |  |  |  | Non-COVID Febrile patients |  |  |  |  |
| --- | --- | --- | --- | --- | --- | --- | --- | --- | --- | --- |
|  | N | TCI | SBI | TDI | CTCI | N | TCI | SBI | TDI | CTCI |
| CRP | 85 | -0.021 | 0.073 | 0.171 | 0.078 | 109 | 0.130 | 0.194* | 0.224* | 0.209* |
| WBC | 89 | 0.258* | 0.301** | 0.349** | 0.332** | 109 | -0.055 | 0.033 | 0.044 | 0.005 |
| Lymphocytes | 89 | -0.063 | -0.107 | -0.124 | -0.107 | 109 | 0.013 | -0.024 | -0.04 | -0.018 |
| Hemoglobin | 89 | -0.118 | -0.056 | -0.103 | -0.103 | 109 | -0.033 | -0.168 | -0.174 | -0.139 |
| BPC | 89 | -0.194 | -0.029 | -0.039 | -0.099 | 109 | 0.001 | -0.01 | 0.016 | 0.003 |
| Neutrophils | 89 | 0.266* | 0.308** | 0.347** | 0.337** | 109 | -0.062 | 0.045 | 0.059 | 0.012 |
| NLR | 89 | 0.287** | 0.258* | 0.294** | 0.308** | 109 | -0.087 | 0.031 | 0.062 | -0.002 |
| PLR | 89 | -0.005 | 0.059 | 0.085 | 0.05 | 109 | -0.114 | -0.037 | -0.016 | -0.067 |

CRP: C-reactive protein, WBC: white blood cell, BPC: blood platelet count, NLR: neutrophil-lymphocyte ratio, PLR: platelet-lymphocyte ratio; \* Correlation is significant at the 0.05 level (2-tailed), \*\* Correlation is significant at the 0.01 level (2-tailed).

### Time course analysis of tongue coating indexes in the COVID group

We also conducted subgroup analysis for COVID patients based on whether they were discharged for home care upon recovery or transferred to another hospital for more intensive care(Figure 3). For each patient, we calculated the initial scores from the first tongue image taken during hospitalization and the last tongue image taken at the end of the hospital stay. As shown in Table 5, initial scores and final scores were compared for patients in the discharged group and the transferred group. For the discharged group, TCI decreased towards the end, with the other indexes showing no significant change. In contrast, for the transferred group, all indexes except the TDI increased before the patient was transferred for more intensive care (Table 5), indicating that tongue coating indexes change according to the direction of disease development.

**Table 5.** Coating indexes at the beginning and towards the end of hospitalization.

| Indexes |  | Discharged group (N=19) |  | Transferred group (N=13) |  |
| --- | --- | --- | --- | --- | --- |
| | | Mean ± SEM | *P* | Mean ± SEM | *P* |
| TCI | Initial score | 8.26 ± 0.64 | 0.026* | 6.92 ± 0.92 | 0.005** |
| | Final score | 6.42 ± 0.86 | | 9.31 ± 0.79 | |
| SBI | Initial score | 6.37 ± 0.52 | 0.236 | 5.31 ± 0.87 | 0.022* |
| | Final score | 5.21 ± 0.78 | | 6.69 ± 0.79 | |
| TDI | Initial score | 4.57 ± 0.79 | 0.704 | 1.92 ± 0.69 | 0.225 |
| | Final score | 4.05 ± 0.98 | | 3.31 ± 1.03 | |
| CTCI | Initial score | 19.21 ± 1.51 | 0.167 | 14.15 ± 2.04 | 0.007** |
| | Final score | 15.68 ± 1.98 | | 19.31 ± 1.55 | |

Data are presented as Mean ± SEM. \**P*<0.05, \*\**P*<0.01 (Student's *t* test)

**Figure 3.** A. Typical examples of tongue images from COVID-19 patients in the Discharged Group and in the Transferred Group. B. Histograms showing the same time sequence of tongue coating indexes of subjects from A. X axis indicates the date of hospital admission.

## Discussion

Using smartphone images of the tongue, this study quantified changes in serval pathological features of tongue coating associated with COVID-19 infection using a comparative approach, controlling for common confounding factors such as acute inflammation, age and diet. We found a significant increase in the appearance of thick, slimy and discolored tongue coating in COVID-19 patients, compared to normal subjects. However, we found no significant difference in all the tongue coating features studied between the COVID and the non-COVID febrile groups. In the longitudinal analysis of COVID-19 patients, we found a reduction in the thickness of tongue coating when the patient recovered from the disease, and increases in coating thickness and slimy biofilm upon further progression of the disease.

Normal tongue coating is a whitish layer, which consists of desquamated epithelial cells, blood cells, metabolites and bacteria adherent to the dorsum of the tongue [14,15]. Increase in coating thickness has been attributed to epithelial cell hyperproliferation with reduced apoptosis and differentiation [16]. Hypothetically, SARS-CoV-2 virus may cause changes in the expression levels of the genes coding related to apoptosis of epithelial cells, causing the accumulation of oral epithelial cells and lead to an increase in coating thickness. In this regard, it is interesting to note that in patients with chronic hepatitis, the level of human epithelial growth factor (EGF) in both saliva and the blood were significantly higher in the thick coating group, corresponding to higher viral load in the liver tissue, compared with the normal coating group [17]. In patients with acute cerebral infarction, thick coating is associated with higher neutrophil count and elevated level of fibrinogen in the blood, compared to patients with normal coating [18]. These studies suggest that the thickness of tongue coating could be associated with viral load and/or inflammation. Our findings that compared with normal subjects, COVID-19 patients have significantly thicker coating, which is correlated with biomarkers of inflammation, and that there is a correlation between coating thickness and disease recovery, support the role of tongue coating thickness in indicating the level of inflammatory response in COVID-19.

The formation of slimy biofilm, corresponding to greasy tongue fur, is a complex phenomenon. Both mucins secreted by salivary glands and epithelial cells, and polysaccharides synthesized by certain microbial cells, may play significant roles in the formation of slimy biofilm [19, 20]. Over production of mucin is a defense response to external stimuli, likely to happen when the oral mucosa is attached by virus directly [4]. However, previous report [5] that COVID-19 patients had more incidents of greasy tongue coating was not substantiated by the current study using non-COVID febrile subjects as control. On the other hand, greasy tongue fur had been reported to be associated with elevated blood level of fibrinogen in stroke patients [18], and with enhanced activity of glossal epithelial cells and increased vascular permeability in rats [21]. The present study found that there was a correlation between the levels of WBC and neutrophils and the level of SBI in COVID-19 patients. In contrast, in non-COVID febrile patients, SBI was correlated with the level of CRP. The reason for such difference in correlation is unclear. It is important to note that oral candidiasis, often caused by compromised immune function, is closely associated with the presence of slimy biofilm on the tongue [22]. Also worth noting are the possibilities that the two groups of patients might be at different stage of their infection and likely to have a different profile in their immune reaction, and that the slimy biofilm is susceptible to oral hygiene and tongue brushing [23]. The myriad of possible factors influencing the appearance of slimy biofilm call for cautions in the interpretation of current results regarding SBI, and further studies are needed to better understand the relationship between COVID-19 and slimy biofilm formation.

Tongue fur discoloration may be caused by several factors. Yellow tongue coating was found to be associated with tongue coating microbiome [24], and it is also susceptible to oral hygiene and smoking [23,25]. Compared with normal subjects, tongue epithelial apoptosis was significantly reduced, and the total number of bacteria was increased in subjects with yellow coating [16]. In patients with acute cerebral infarction, thick and yellow tongue coating was associated with higher neutrophil count and elevated level of fibrinogen [18]. In the current findings, TDI had the highest correlation coefficients with the inflammatory biomarkers WBC and neutrophils, compared with other coating indexes of COVID-19 patients. Therefore, tongue fur discoloration may be indicative for the level of inflammatory response in COVID-19 patients.

Taken together, notwithstanding the complex mechanisms associated with various pathological coating features, a clear picture emerging from the current study is the association between pathological tongue coating and inflammatory responses in COVID-19, as indicated in the correlation analysis between coating indexes and inflammatory biomarkers. Although the correlation is weak in general, they are consistent across several inflammatory biomarkers and pathological coating features. The reason for a different pattern of correlation between pathological tongue coating and inflammatory biomarkers seen in the non-COVID febrile group may be due to different nature of the associated inflammatory and immune responses [26].

To our knowledge, this is the first COVID tongue study that has employed a quantitative method and has controlled for important confounding factors influencing tongue coating [5, 27, 28]. Our findings are consistent with the results of many other previous studies, which have showed that thick, slimy and yellow tongue coating are characteristics of febrile subjects [5, 29]. Also in agreement with previous reports are the findings that tongue thickness coating reduces as patients recovering from COVID-19 [5, 30]. Previous studies suggested that slimy biofilm was a significant characteristic of patients with COVID-19 [5, 30]. While we could not find any difference in the level of slimy biofilm between COVID and non-COVID febrile subjects, we did find that COVID patients with worsening condition showed increased levels of thick and slimy coating, compared with their own tongue coating at the beginning of COVID infection. Furthermore, we have demonstrated a specific correlation pattern between pathological tongue coating and inflammatory biomarkers in COVID-19. These findings are of practical interest, suggesting that tongue coating changes could be used as one of the indicators for the direction of disease development in COVID-19 patients. Specifically, reduction in coating thickness may indicate recovery and persistent increase in coating thickness and/or slimy biofilm coverage may signify deterioration.

There are many strengths of the current study. Firstly, this study controlled for major confounding factors known to influence tongue coating, including acute infection, age and diet. Second, it adopted quantitative methods for tongue coating assessment, allowing for correlation analysis with inflammatory biomarkers. Last but not the least, it went through a lengthy process of evaluating rater consistency and training of assessors, and used blinding for tongue coating assessment to reduce assessor bias. Taken together, the current study is underpinned by vigorous methodological design and its results are expected to have good validity in the real world.

There are a number of limitations in this study that are worth noting. For example, the source of COVID-19 patients are all from Wuhan at a particular time period. Thus, it is not known whether new COVID variants could produce specific tongue coating patterns not seen in the current study. Due to the lack of matching subjects, we have to exclude many older age COVID-19 patients, resulting in a relatively small sample size. As well, because the medical records of COVID-19 patients are incomplete, it is not possible to control for any comorbid disease in the study population. Therefore, findings from the current study should be interpreted with the above limitation in mind.

In conclusion, COVID-19 patients have pathological tongue coating that is associated with inflammatory responses, and their coating changes may indicate the direction of disease development. Further studies are warranted to explore the application of tongue imaging as an adjunct diagnostic method in monitoring the development of COVID-19.

## Data Availability

Data cannot be shared publicly because of patient privacy. Data are available from corresponding author in Hong Kong Baptist University for researchers who meet the criteria for access to confidential data.

## Acknowledgements

We thank Dr Fan Mingcong and staff of Wuhan Xiao Pan Jing Hotel Management Co. Ltd for contributing tongue images.

## Compliance with ethics guidelines

Zhi Chun Wang, Xi Hong Cai, Jeremy Chan, Yi Yi Chan, Xiaotong Chen, Ching Wan Cheng, Donghui Huang, Bei-ni Lao, Xu-sheng Liu, Aiping Lu, Huixian Wang, Helen Zhang, Xuebin Zhang, Shi Ping Zhang declare that they have no conflict of interest.

Given the retrospective nature of this study, informed consent was waived in the collection of tongue images from COVID-19 subjects.

The study was approved by the Ethics Committee of Hong Kong Baptist University (REC/19-20/0400), Ethics Committee of The Third Affiliated Hospital of Guangzhou University of Chinese Medicine (AF/SC-07/02.0) and Ethics Committee of Guangdong Provincial Hospital of Chinese Medicine (AF/04-06.1/10.0).

